# Worries and concerns among healthcare workers during the coronavirus 2019 pandemic: a web-based cross-sectional survey

**DOI:** 10.1101/2020.06.09.20126045

**Authors:** Yuki Sahashi, Hirohisa Endo, Tadafumi Sugimoto, Takeru Nabeta, Kimitaka Nishizaki, Atsushi Kikuchi, Shingo Matsumoto, Hiroyuki Sato, Tadahiro Goto, Kohei Hasegawa, Yuya Matsue

## Abstract

**Background:** Healthcare workers (HCWs) treating and caring for patients with emerging infectious diseases often experience psychological distress. However, the psychological impact and behavior change of the coronavirus disease 2019 (COVID-19) pandemic among HCWs are still unknown. This study aimed to investigate the worries and concerns of HCWs regarding the COVID-19 pandemic.

**Methods:** In this cross-sectional survey, a web-based questionnaire was distributed among HCWs working in hospitals or clinics across Japanese medical facilities from April 20 to May 1, 2020. The questionnaire comprised items on demographics, worries and concerns, perceptions regarding the sufficiency of information, and behavioral changes pertaining to the COVID-19 pandemic.

**Results:** A total of 4386 HCWs completed the survey; 1648 (64.7%) were aged 30-39 years, 2379 (54.2%) were male, and 782 (18.1%) were frontline HCWs, directly caring for patients with COVID-19 on a daily basis. 3500 HCWs (79.8%) indicated that they were seriously worried about the pandemic. The most frequent concern was the consequence of becoming infected on their family, work, and society (87.4%). Additionally, the majority (55.5%) had restricted social contact and almost all HCWs endorsed a shortage in personal protective equipment (median, 8/9 (interquartile range; 7-9) on a Likert scale). There was no significant difference in the degree of worry between frontline and non-frontline HCWs (8/9 (7-9) vs. 8/9 (7-9), p=0.25). Frontline HCWs, compared to non-frontline HCWs, were more likely to have the need to avoid contact with families and friends (24.8% vs. 17.8%, p<0.001) and indicated that they cannot evade their professional duty during the COVID-19 pandemic (9/9 (7-9) vs. 8/9 (6-9), p<0.001). Further, the extremely low proportion of frontline HCWs reported that they would take a leave of absence to avoid infection (1.2%).

**Conclusions:** Both frontline and non-frontline HCWs expressed comparable concerns regarding the COVID-19 pandemic. Because HCWs, especially frontline HCWs, reported that they cannot be obliged to do avoid their duty, effective mental health protection strategies should be developed and implemented for HCWs.

## Background

Since the first case of severe acute respiratory syndrome coronavirus-2 (SARS-CoV-2) infection was reported in the Wuhan province of China at the end of 2019, the number of confirmed cases and deaths has been increasing worldwide. As of May 1, 2020, the number of patients with coronavirus disease 2019 (COVID-19) has reached over 3 million, and more than 220,000 deaths have been confirmed.(1)

After the outbreak of COVID-19, a large number of healthcare workers (HCWs) became infected with SARS-CoV-2, accounting for 4%–11% of confirmed cases.(2, 3) In the context of this unprecedented pandemic, frontline HCWs, who have direct exposures to patients with COVID-19 on a daily basis, are at high risk of developing mental health problems due to concerns regarding the COVID-19 pandemic. COVID-19 pandemic-associated mental distress is attracting considerable attention from the mental health community and the general public, as it has already become a notable problem for frontline HCWs at the epicenter of the COVID-19 pandemic.(4, 5) Despite these clinical research importance, HCWs, whether frontline or non-frontline, are at risk of infection and may be exposed to significant psychological distress.

To address the knowledge gap in the literature, this study aimed to investigate the psychological distress of HCWs regarding the COVID-19 pandemic. Understanding the psychological impact of the COVID-19 pandemic on HCWs should help in providing HCWs with safe and optimal working conditions, and may prevent the healthcare system from becoming overwhelmed.

## Methods

### Study design and setting

This cross-sectional, web-based survey was carried out from April 20, 2020 to May 1, 2020. The target participants of this survey were HCWs in Japanese hospitals and clinics who were—directly or indirectly—treating patients with COVID-19. This study was approved by the ethical committee of Juntendo University (No. 2020025) and was performed in accordance with the ethical principles of the Declaration of Helsinki. All authors take complete responsibility for the integrity of the survey and study design, data collection, and the accuracy of the data analysis. The requirement for written informed consent was waived because of the nature of study design. Instead of providing signed written informed consent, responders who gave a consent to participate in this study did so by filling in the agreement portion of the survey form.

### Study participants and recruitment

The participating HCWs included physicians, nurses, pharmacists, radiology technicians, clinical engineers, and physical therapists. We classified participants caring for patients with COVID-19 on a daily basis as frontline HCWs, specifically those who answered, “I am currently doing it routinely” to the question “Are you currently caring for patients with COVID-19”. (see the **Supplemental Appendix, Q34**). The online survey was distributed to HCWs through email lists of hospital or local medical associations, medical school alumni associations, and closed medical groups on social media. The questionnaire and aim of the study were sent to each member of the medical group, along with information that participation in the survey was voluntary. The web-based questionnaire was distributed on April 20, 2020, with a predefined closure date of May 1, 2020.

Responders could refuse to give a consent to participate in the study by simply ticking a checkbox at the end of the questionnaire; data from such responders were excluded from the analysis.

### Survey items

The web-based survey included 34 items according to a previous study for H1N1 influenza pandemic.(6) The survey was comprised two parts. The first part collected data on the participants’ demographics and characteristics, including age, sex, type of occupation, the prefecture in which they lived, the department in which they worked, clinical experience with COVID-19, type of hospital (infectious disease-designated medical institution or not), number of years of practice, and whether they lived with any family members or children.

The second part of the survey comprised three sections comprising 23 items examining (1) their worries and concerns (the degree and content of their worries, their concern regarding the risk of being infected with SARS-CoV-2, the insufficiency of PPE in their facility); (2) perceptions regarding the availability and need of information on COVID-19 (perceived sufficiency of information about COVID-19 symptoms, treatment, transmission routes, and preventive measures, whether their facility provided clear information on COVID-19, how much information about an infectious disease the respondent would prefer to have); and (3) their behaviors during the COVID-19 pandemic (intentional behavior changes, such as restricted social contact, work avoidance, and their sense of duty). The survey forms are shown in the **Supplemental Appendix**.

Most items were dichotomous (yes/no) or scored on a 9-point Likert scale ranging from 1 to 9, corresponding to ‘very little’ (strongly disagree, very low) and ‘very much’ (strongly agree, very high), respectively. Some items were presented as multiple-choice questions (see the **Supplemental Appendix**). The questionnaire was anonymous, and the privacy policy of the individual’s posted information was noted.

### Statistical analysis

We summarized the data according to frontline and non-frontline HCWs. For the participant characteristics, the continuous variables are expressed as the mean ± standard deviation (SD) or median with interquartile range (IQR), depending on the distribution of the data. The categorical variables are expressed as percentages. The continuous variables were compared using the one-way analysis of variance or the Mann-Whitney U test; the categorical variables were compared using the chi-square test. Responses on a 9-point Likert scale were not analyzed as ordinal variables, but as continuous variables.(7, 8) The Cronbach alpha of reliability for these data was 0.71. The number of cumulative and new COVID-19 cases per 1 million people in the respondent’s prefecture on the day of survey completion was obtained from data published by Japan’s Ministry of Health, Labour, and Welfare (9). We defined the epidemic area as the top 10 regions among 47 prefectures in terms of the cumulative number of patients with COVID-19 per 1 million people. In accordance with the recent report by Idogawa *et al. (10)*, we also calculated the *k*-value as an indication of infection propagation activity.(11) The *k*-value was calculated with the formula *y* = exp (*kt* + *C*), using the values of the prior 3 days. P-values of <0.05 were considered statistically significant. All data were analyzed using JMP version 12.2 for Windows (SAS Institute, Cary, NC).

## Results

### Participants’ characteristics

Participant characteristics are summarized in **Table 1**. Among 4419 responders, 33 who declined to participate in the study were excluded; the analytic cohort consisted of 4386 participants. Of these, many participants were 30-39 years old, there were 2379 men (54.2%), 1365 (31.1%) physicians working in hospitals, 338 (7.7%) general practitioners, 1173 (26.7%) nurses (hospital and clinic nurses), 246 (5.6%) pharmacists, 357 (8.1%) radiology technicians, 107 (2.4%) clinical engineers, and 800 (18.2%) physical therapists. Tokyo was the most frequently indicated region of residence (1412[32.2%]), and 1361(31.0%) participants worked in infectious disease-designated medical institutions. Many participants lived with family members (3196 [72.9%]), and approximately a half had children (2188 [49.9%]). At the time of survey completion, the mean cumulative number of patients with a positive polymerase chain reaction test for SARS-CoV-2 in the respondent’s prefecture was 106 patients per one million people.

**Table 1.** Baseline characteristics of study participants (n=4386)

| Variables | Entire cohort<br>(n=4386) | HCW's role |  | P-value |
| --- | --- | --- | --- | --- |
|  |  | Frontline <sup>a</sup><br>(n=782, 18.2%) <sup>b</sup> | Non-frontline<br>(n=3516, 81.8%) <sup>b</sup> |  |
| Age, years | 37 ± 11 | 38 ± 10 | 38 ± 11 | 0.16 |
| Male, n (%) | 2379 (54.2) | 490 (62.7) | 1838 (52.3) | <0.001 |
| Speciality, n (%) |  |  |  | <0.001 |
| Hospital physician | 1365 (31.1) | 298 (38.1) | 1048 (29.8) |  |
| General practitioner | 338 (7.7) | 30 (3.8) | 304 (8.7) |  |
| Hospital nurse | 929 (21.1) | 161 (20.6) | 750 (21.4) |  |
| Clinic nurse | 244 (5.6) | 56 (7.2) | 181 (5.2) |  |
| Pharmacist | 246 (5.6) | 26 (3.3) | 217 (6.2) |  |
| Radiology technician | 357 (8.1) | 114 (14.6) | 231 (6.6) |  |
| Clinical engineer | 107 (2.4) | 32 (4.1) | 73 (2.1) |  |
| Physical therapist | 800 (18.2) | 65 (8.3) | 711 (20.2) |  |
| Infectious disease-designated medical institution, n (%) | 1361 (31.0) | 321 (41.1) | 1017 (28.9) | <0.001 |
| Specialty (physician, nurse), n (%) |  |  |  | <0.001 |
| Physician | 1181 (39.4) | 220 (38.8) | 941 (39.6) |  |
| Intensivist/emergency physician | 570 (19.0) | 232 (40.9) | 326 (13.7) |  |
| Surgeon | 423 (14.1) | 23 (4.1) | 394 (16.6) |  |
| Others | 820 (27.4) | 92 (16.2) | 717 (30.2) |  |
| Epidemic area | 2604 (59.4) | 499 (63.8) | 2054 (58.4) | 0.006 |
| k-value | 0.034 (0.02-0.037) | 0.035 (0.021-0.037) | 0.034 (0.02-0.037) | 0.03 |
| Cumulative number of patients with COVID-19 in the region of residence (per million) | 106 (70-237) | 110 (87-237) | 106 (68-237) | 0.003 |
| Main workspace, n (%) |  |  |  |  |
| Outpatient | 1959 (44.7) | 296 (37.9) | 1620 (46.1) | <0.001 |
| Ward | 2361 (53.8) | 387 (49.5) | 1928 (54.8) | 0.007 |
| Emergency department | 988 (22.5) | 323 (41.3) | 643 (18.3) | <0.001 |
| Intensive care unit | 951 (21.7) | 325 (41.6) | 607 (17.3) | <0.001 |
| Operation room | 711 (16.2) | 115 (14.7) | 582 (16.6) | 0.21 |
| Others | 947 (21.6) | 160 (20.5) | 757 (21.5) | 0.51 |
| Live alone, n (%) | 1190 (27.1) | 219 (28.0) | 949 (27.0) | 0.57 |
| Have children, n (%) | 2188 (49.9) | 400 (51.2) | 1747 (49.7) | 0.46 |
| Infected with SARS-CoV-2, n (%) | 14 (0.3) | 6 (0.8) | 8 (0.2) | 0.02 |
| Contact with patients with COVID-19, n (%) | 862 (19.7) | 461 (59.0) | 381 (10.8) | <0.001 |
| Family member or colleague infected with SARS-CoV-2, n (%) | 212 (4.8) | 70 (9.0) | 139 (4.0) | <0.001 |

COVID-19, coronavirus disease 2019; epidemic area, the top 10 regions among 47 prefectures in terms of the cumulative number of COVID-19 cases per 1 million people; HCW, health care worker. The data are presented in n (%) otherwise being specified.
<sup>a</sup> Frontline HCWs is defined as participants caring for patients with COVID-19 on a daily basis, specifically those who answered, "I am currently doing it routinely" to the question "Are you currently caring for patients with COVID-19".
<sup>b</sup> 88 participants did not respond the question "Are you currently caring for patients with COVID-19".

Additionally, 728 participants (18.2%) were classified as frontline HCWs. In this survey, frontline HCWs were more likely to be male (62.7 %), work at infectious disease-designated medical institutions (41.1%), and work in an intensive care unit or emergency department (40.9%) than were non-frontline HCWs. In contrast, non-frontline HCWs were more likely to be general practitioner or surgeons than were frontline HCWs. Frontline HCWs were more likely to have been infected with SARS-CoV-2 compared with non-frontline HCWs (0.8% vs. 0.2%, p<0.001). In addition, their family members and colleagues were also more likely to have been infected with SARS-CoV-2 compared with those of non-frontline HCWs (9.0% vs. 4.0%, p<0.001).

### Worries and concerns about the COVID-19 pandemic

The degree and detailed content of the respondents’ worries and concerns regarding the COVID-19 pandemic are shown in **Table 2**. Almost all (98.6%) respondents were worried about the COVID-19 pandemic, with a high degree of worry (score of 7-9) in most (79.8%) respondents. The most frequent concern was the impact that becoming infected would have on their family, work, and society (87.4%), followed by the risk of family members and relatives becoming infected by SARS-CoV-2 (84.2%).

**Table 2.** Healthcare workers’ worries and concerns about the COVID-19 pandemic

| Variables | Entire cohort<br>(n=4386) | HCW's role |  | P-value |
| --- | --- | --- | --- | --- |
|  |  | Frontline <sup>a</sup><br>(n=782, 18.2%) <sup>b</sup> | Non-Frontline<br>(n=3516, 81.8%) <sup>b</sup> |  |
| Worried about the COVID-19 pandemic, Yes, n (%) | 4324 (98.6) | 770 (98.5) | 3466 (95.6) | 0.81 |
| Degree of worry <sup>c</sup> | 8 (7-9) | 8 (7-9) | 8 (7-9) | 0.25 |
| I mostly worry about, n (%) <sup>d</sup> |  |  |  |  |
| The consequences on my functional ability | 3807 (87.4) | 660 (84.9) | 3071 (88.0) | 0.02 |
| The risk of infecting family members or other relatives | 3667 (84.2) | 650 (83.7) | 2936 (84.1) | 0.75 |
| The disease's dangerousness | 3136 (72.0) | 550 (70.8) | 2516 (72.1) | 0.47 |
| Isolation from family and/or the social environment | 1304 (29.9) | 238 (30.6) | 1035 (29.7) | 0.59 |
| Perceived risk of being infected by SARS-CoV-2* | 6 (5-7) | 7 (5-8) | 6 (5-7) | <0.001 |
| I think that being infected with SARS-CoV-2 would have major consequences to my health | 7 (6-9) | 7 (6-9) | 7 (6-9) | 0.18 |
| I believe that the infection is difficult to treat | 7 (5-8) | 7 (5-8) | 7 (5-8) | 0.001 |
| I think that my department has been well prepared for the COVID-19 pandemic | 5 (3-7) | 5 (4-7) | 5 (3-6) | <0.001 |
| I think that it is important to have a service offering psychological support regarding my concerns about the pandemic | 6 (6-8) | 6 (6-8) | 6 (6-7) | 0.07 |
| I believe that the recommended preventive measures are effective | 6 (5-7) | 7 (5-7) | 6 (5-7) | <0.001 |
| I think that the required personal protective equipment are not unavailable | 8 (7-9) | 9 (7-9) | 8 (7-9) | 0.01 |

COVID-19, coronavirus disease 2019; HCW, healthcare worker; IQR, interquartile range; SARS-CoV-2, severe acute respiratory syndrome coronavirus-2/
<sup>a</sup> Frontline HCWs is defined as participants caring for patients with COVID-19 on a daily basis, specifically those who answered, "I am currently doing it routinely" to the question "Are you currently caring for patients with COVID-19".
<sup>b</sup> 88 participants did not respond the question "Are you currently caring for patients with COVID-19".
<sup>c</sup> The value of 9-point Likert scale is presented in median value (IQR). 1 = strongly disagree (\*very low), 9 = strongly agree (\*very high).
<sup>d</sup> Only 4324 participants who responded "yes" to the question "Are you worried about the COVID-19 pandemic"

There were no significant differences between frontline HCWs and non-frontline HCWs in their concern about the health risk of the disease itself (70.8% vs. 72.1%, p=0.47), the risk of infection in family members or other relatives (83.7% vs. 84.1%, p=0.75), and isolation from family and/or the social environment (30.6% vs. 29.7%, p=0.59).

Overall, respondents rated the degree of sufficiency of their department’s preparation for the COVID-19 pandemic as relatively low (5/9, IQR; 3-7). However, frontline HCWs were more likely to indicate that their department has been well prepared for the COVID-19 pandemic than were non-frontline HCWs (5/9, IQR; 4-7 vs. 5/9, IQR; 3-6, p<0.001). Both frontline and non-frontline HCWs reported that the availability of PPE was greatly insufficient (8/9, IQR; 7-9).

### Perceptions regarding the sufficiency of information on COVID-19

**Table 3** summarizes the responses to the sufficiency of information on COVID-19. Although there was a wide variation in responses, the perceived sufficiency of available information on the symptoms, treatment, transmission routes, and preventive measures of COVID-19 was rated as relatively low (**Figure 1**). More frontline HCWs, compared to non-frontline HCWs, reported that they had sufficient information about COVID-19 health issues and their department provided adequate information. Moreover, approximately half of the participants (2117 [48.3%] indicated that they wish to have as much information as possible.

**Table 3.** Healthcare workers’ perceptions regarding the sufficiency of information about COVID-19 and general health information needs

| Variables | Entire cohort<br>(n=4386) | HCW's role |  | P-value |
| --- | --- | --- | --- | --- |
|  |  | Frontline <sup>a</sup><br>(n=782, 18.2%) <sup>b</sup> | Non-Frontline<br>(n=3516, 81.8%) <sup>b</sup> |  |
| I believe that I have had sufficient information about <sup>c</sup> |  |  |  |  |
| COVID-19 symptoms, median (IQR) | 6 (3-7) | 6 (4-7) | 5 (3-7) | <0.001 |
| COVID-19 treatment, median (IQR) | 4 (2-6) | 5 (3-7) | 4 (2-6) | <0.001 |
| COVID-19 transmission routes, median (IQR) | 5 (3-7) | 6 (3-7) | 5 (3-7) | <0.001 |
| COVID-19 preventive measures, median (IQR) | 6 (3-7) | 6 (4-8) | 6 (3-7) | <0.001 |
| I believe that my department has provided adequate information about the COVID-19 pandemic, median (IQR) <sup>c</sup> | 6 (4-7) | 6 (5-8) | 6 (4-7) | <0.001 |
| General health-information needs, median (IQR) <sup>d</sup> | 4 (3-5) | 5 (3-5) | 4 (3-5) | 0.39 |

COVID-19, coronavirus disease 2019; HCW, healthcare worker; IQR, interquartile range.
<sup>a</sup> Frontline HCWs is defined as participants caring for patients with COVID-19 on a daily basis, specifically those who answered, "I am currently doing it routinely" to the question "Are you currently caring for patients with COVID-19".
<sup>b</sup> 88 participants did not respond the question "Are you currently caring for patients with COVID-19".
<sup>c</sup> The value of 9-point Likert scale is presented in median value (IQR). 1 = strongly disagree, 9 = strongly agree.
<sup>d</sup> 5-point Likert scale: 1 = for a disease that I might suffer, I prefer having no more information than needed, 5 = I prefer to have as much information as possible

**Figure 1.**
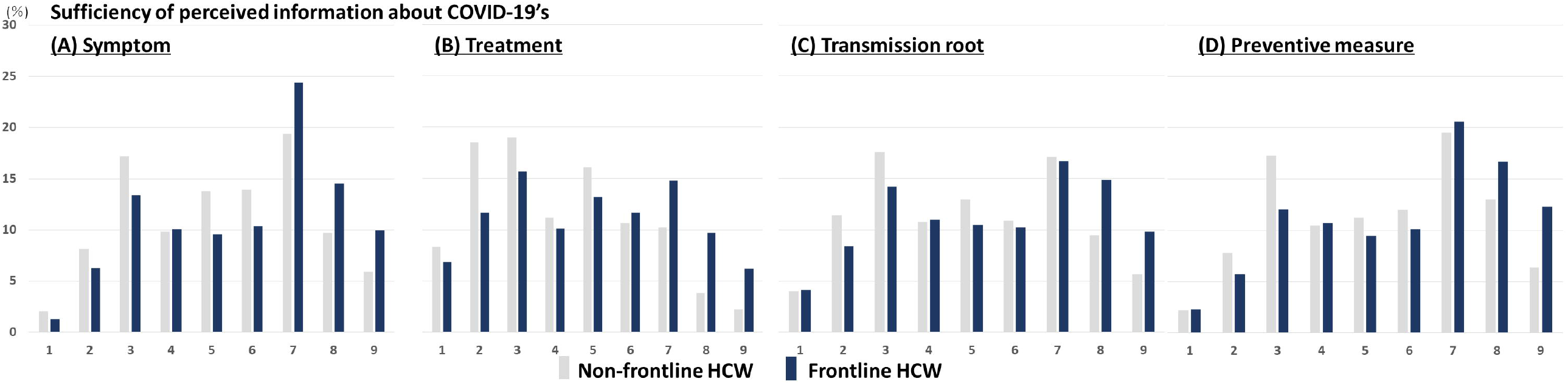
Sufficiency of perceived information about COVID-19’s. The perceived degree of sufficiency of information regarding COVID-19’s (A) symptoms, (B) treatment, (C) route of transmission, and (D) preventive measures. The x axis represents the response on a 9-point Likert scale, with 1 point indicating that the respondent felt strongly that information was lacking and 9 points indicating that they felt strongly that the information was sufficient. For all 4 issues, frontline HCWs are more satisfied with the amount of information than were non-frontline HCWs.

### Behavioral changes during the COVID-19 pandemic

As shown in **Table 4**, the majority (2434 [55.5%]) indicated that they had restricted social contact because of their risk of contracting SARS-CoV-2. This was more common among frontline HCWs than among non-frontline HCWs (64.2% vs. 53.6%, p<0.001). Additionally, 838 HCWs (19.1%) indicated that they felt shunned by their family members and friends. Only 94 HCWs (2.1%) indicated that they would take a leave of absence due to COVID-19 worries and concerns. Although almost all HCWs (98.6%) indicated that they were concerned about the COVID-19 pandemic, 3246 HCWs (74.0%) indicated that it was highly impossible (7-9 points on the Likert scale) to evade their duties in the public emergency. Furthermore, frontline HCWs were more likely to report the need to avoid contact with families and friends (24.8% vs. 17.8%, p<0.001) and believed that it was highly impossible to leave their work during the COVID-19 pandemic (9/9, IQR; 7-9 vs. 8/9, IQR; 6-9, p<0.001).

**Table 4.** Intentional behavioral changes associated with worry, as well as the degree of worry, about the COVID-19 pandemic

| Variables | Entire cohort<br>(n=4386) | HCW's role |  | p-value |
| --- | --- | --- | --- | --- |
|  |  | Frontline <sup>a</sup><br>(n=782, 18.2%) <sup>b</sup> | Non-Frontline<br>(n=3516, 81.8%) <sup>b</sup> |  |
| I have restricted my social contacts because my work environment is considered "dangerous". Yes, n (%) | 2434 (55.5) | 502 (64.2) | 1886 (53.6) | <0.001 |
| I feel that my family members and friends avoid contact with me, because I work in a "high-risk" environment. Yes, n (%) | 838 (19.1) | 194 (24.8) | 626 (17.8) | <0.001 |
| Lately I have been so concerned about COVID-19 that I would take a leave to avoid going to work. Yes, n (%) | 94 (2.1) | 9 (1.2) | 82 (2.3) | 0.03 |
| In a COVID-19-related emergency situation, how possible would it be to avoid your duties? <sup>c</sup> | 8 (6-9) | 9 (7-9) | 8 (6-9) | <0.001 |

COVID-19, coronavirus disease 2019; HCW, healthcare worker; IQR, interquartile range.
<sup>a</sup> Frontline HCWs is defined as participants caring for patients with COVID-19 on a daily basis, specifically those who answered, "I am currently doing it routinely" to the question "Are you currently caring for patients with COVID-19".
<sup>b</sup> 88 participants did not respond the question "Are you currently caring for patients with COVID-19".
<sup>c</sup> The value of 9-point Likert scale is presented in median value (IQR). 1 = highly possible, 9 = not at all possible

## Discussion

In this large survey of 4386 HCWs across Japan, we found 1) 98.6% indicated that they are very worried about the COVID-19 pandemic; 2) HCWs, regardless of frontline or non-frontline workers, indicated that the available information on COVID-19 is insufficient, and that they wish to have as much information as possible; 3) the majority of HCWs, especially frontline HCWs, indicated that it is impossible to evade their duties, despite a lack of sufficient information and PPE. Our findings highlight the psychological distress of HCWs engaged in their work with great responsibility and a lack of information amid the public health emergency of COVID-19.

The mental distress of HCWs during infectious disease pandemics has been previously described, especially for the 2003 SARS and 2009 H1N1 influenza pandemics.(6, 12) Compared to the present study on the COVID-19 pandemic, in the 2009 H1N1 influenza pandemic, a smaller proportion of HCWs (56.7%) indicated that they were worried about the disease.(6) In the present study, the HCWs reported more fear and worry compared to that in previous studies on the mental health of HCWs during the emergence of other infectious diseases, such as Middle East respiratory syndrome(13) and SARS.(14) One potential reason for these apparent difference in the degree of worry involves the perceived insufficiency of information, as knowing the latest and most accurate health information (e.g., treatment, transmission, and precautions) reduces the impact of a pandemic on anxiety and depression.(15) The degree of satisfaction regarding the sufficiency of available information was lower in the present COVID-19 study than in a previous study on psychological distress in HCWs during the 2009 H1N1 influenza pandemic.(6) Further, frontline HCWs receive a flood of information from various medical societies, social online news media, and colleagues, which can create uncertainty and be overwhelming for many HCWs. Up-to-date and accurate information on COVID-19 should be delivered promptly to HCWs to mitigate stress stemming from uncertainties regarding this disease.

In our survey, there was no significant difference between frontline and non-frontline HCWs in the degree of worry. In other words, the non-frontline HCWs had worries and concerns about COVID-19 as well as the frontline HCWs, which differs from the results of previous studies that revealed frontline HCWs felt more anxious(6, 12, 16). This could be related to SARS-CoV-2’s uncertain transmission route and strong infectivity; patients with COVID-19 can be infective before becoming symptomatic.(17) These characteristics make SARS-CoV-2 different from other viruses. Additionally, studies conducted in China reported that HCWs engaged in COVID-19 treatment on the frontline are more mentally distressed than were those not on the frontline, which is inconsistent with the present results.(5) This may be due to differences in the timing of the survey relative to the start of the pandemic and the number of people infected in the region. In fact, the infection rate in Wuhan (one of the cities in China where the study by Lai et al.(5) was conducted) was dozens of times higher than that in Japan.(1) In addition, information regarding the route of transmission of SARS-CoV-2 was much less clear when the study by Lai et al.(5) was conducted.

In the present study, HCWs generally felt motivated to work during the COVID-19 pandemic, as shown by the extremely low proportion of HCWs who indicated that they would take a leave of absence to avoid infection (2.1%) and the high degree of agreement with the statement that it was impossible to avoid their duties (mean, 7.3 ± 1.9). These values are lower and higher, respectively, than those in a previous study on the H1N1 influenza pandemic (would take a leave of absence to avoid infection: 4.3%: impossible to avoid their duties: 5.4±2.8). (6) This dissociation between the degree of HCWs’ worries and how likely they feel they can avoid their duty should be acknowledged, because this may be one of the main factors affecting the mental health of HCWs, not only during the pandemic, but also after the pandemic. Indeed, it has been reported that HCWs experience a high level of burnout and can suffer from post-traumatic syndrome for a long time.(18, 19) Various studies have suggested that active mental support interventions should be available for healthcare providers in every healthcare situation.(20, 21) This concept is supported by the present study results, as many of the participants indicated that mental support for HCWs would be useful. Given that both frontline and non-frontline HCWs have strong anxiety and believe that mental support is beneficial, long-term active interventions for anxiety due to the COVID-19 pandemic should be considered not only for frontline HCWs, but also for non-frontline HCWs.

The use of PPE is essential in the clinical practice of treating COVID-19, and a shortage of PPE increases the risk of infection in healthcare providers.(22) To cope with a shortage in PPE, research regarding PPE reprocessing methods has been performed in various medical facilities,(23, 24) following the WHO’s proposal for the appropriate use of PPE.(25) As of April-May 2020, Japan has a massive shortage in PPE, similar to that globally,(26) especially in metropolitan areas. The spread of SARS-CoV-2 infection among HCWs exacerbated nosocomial cases of SARS-CoV-2 infection.(27, 28) Consequently, several medical institutions in Japanese epidemic areas ceased to function, and the regional healthcare system was on the verge of collapse. As almost all HCWs endorsed a massive shortage in PPE, which is considered to be a major cause of anxiety among HCWs, a proper discussion on rational PPE use and supply is needed.

## Limitations

The present study has several potential limitations. First, there may be a selection bias. Although the survey was distributed widely, the study sample is not a random sample of all HCWs in Japan. Additionally, a response bias (volunteer effect) should be considered in this setting. Not all HCWs who received this questionnaire responded, including those who were too stressed to respond or were not sufficiently interested in this survey. Because of the study’s design, we were unable to calculate the exact proportion of respondents and characterize the differences between respondents and non-respondents. Second, in the setting of emergent COVID-19 pandemic, the degree of psychological distress was not precisely quantified by widely used and well-validated questionnaires. Yet, we have developed the survey based on the H1N1 influenza pandemic literature. (6) Lastly, our results may have limited generalizability despite the large-scale data collected from diverse settings and geographical regions across Japan. While it is tempting to dismiss the broader applicability, the observed findings are plausible and potentially generalized to other healthcare settings.

## Conclusions

In conclusions, based on the nationwide survey of 4368 HCWs during the COVID-19 pandemic, we found that almost all HCWs continue to work, despite a lack of information and several worries and concerns such as the infection risk of their family or relatives and the consequences on their functional ability. Both frontline and non-frontline HCWs expressed comparable but substantial concerns regarding the COVID-19 pandemic and the serious shortage of PPE. Effective mental health protection strategies to prevent burnout and depression should be developed and implemented for HCWs, who are trying hard to fulfill their responsibilities in tackling the public health crisis.

## Data Availability

All data relevant to the study are included in the article. No additional data available. The data underlying the results of this study are available upon request due to ethical restrictions imposed by the Juntendo University Hospital Institutional Review Board. Due to the sensibility of the data, and in order to ensure full anonymity, confidentiality and data protection for the participants, the full survey data cannot be made accessible to the public. Interested researchers may contact the corresponding author via email at ""

## Acknowledgement

We would like to thank all health care professionals who provide COVID-19 treatment and care throughout Japan and those who answered the questionnaire. We would also like to express our sincere gratitude to all representatives of working groups those who cooperated in the distribution of the questionnaire.

## Author contributions

H.E. had full access to all the data in the study and take responsibility for the integrity of the data and the accuracy of the data analysis. Concept and design: Y.M. Acquisition, analysis and interpretation of the data: Y.S., H.E., T.S.,, T.N., K.N., A.K., S.M., H.S., and Y.M. Drafting of the manuscript: Y.S, and H.E. Critical revision of the manuscript for important intellectual content: T.S., T.N., K.N., A.K., S.M., H.S., T.G., K.H., and Y.M. Statistical analysis: H.E. Supervision: T.G., H.K., and Y.M.

## A funding statement

This research received no specific grant from any funding agency in the public, commercial or not-for-profit sectors

## Conflict of Interest Disclosures

Dr. Yuya Matsue is affiliated with a department endowed by Philips Respironics, ResMed, Teijin Home Healthcare, and Fukuda Denshi; received an honorarium from Otsuka Pharmaceutical Co.; and received consulting fees from Edwards Lifesciences and Bristol-Myers Squibb. The remaining authors have no conflicts of interest to report.

## Notes

### Author Declarations

This study was approved by the ethical committee of Juntendo University (No. 2020025).

